# LMP-TX: An AI-driven Integrated Longitudinal Multi-modal Platform for Early Prognosis of Late Onset Alzheimer’s Disease

**DOI:** 10.1101/2024.10.02.24314019

**Authors:** Victor OK Li, Jacqueline CK Lam, Yang Han

## Abstract

Alzheimer’s Disease (AD) is the 7th leading cause of death worldwide. 95% of AD cases are late-onset Alzheimer’s disease (LOAD), which often takes decades to evolve and become symptomatic. Early prognosis of LOAD is critical for timely intervention before irreversible brain damage. This study proposes an Artificial Intelligence (AI)-driven longitudinal multi-modal platform with time-series transformer (LMP-TX) for the early prognosis of LOAD. It has two versions: LMP-TX utilizes full multi-modal data to provide more accurate prediction, while a lightweight version, LMP-TX-CL, only uses simple multi-modal and cognitive-linguistic (CL) data. Results on prognosis accuracy based on the AUC scores for subjects progressing from normal control (NC) to early mild cognitive impairment (*e*MCI) and *e*MCI to late MCI (*l*MCI) is respectively 89% maximum (predicted by LMP-TX) and 81% maximum (predicted by LMP-TX-CL). Moreover, results on the top biomarkers predicting different states of LOAD onsets have revealed key multi-modal (including CL-based) biomarkers indicative of early-stage LOAD progressions. Future work will develop a more fine-grained LMP-TX based on disease progression scores and identify the key multi-modal and CL-based biomarkers predictive of fast AD progression rates at early stages.

## 1. Introduction

Alzheimer’s Disease (AD) is the 7th leading cause of death worldwide. 95% of AD cases occur after age 65, i.e. Late Onset Alzheimer’s Disease (LOAD). However, LOAD often takes decades to evolve and become symptomatic, making it challenging to provide timely intervention at early stages. Statistical methods and artificial intelligence (AI) approaches have been proposed for LOAD diagnosis (how likely one will get LOAD onsets now) and prognosis (how likely one will get LOAD onsets in the future). Statistical models aim to characterize long-term disease timelines by describing a set of biomarker trajectories (often under certain assumptions, e.g., the trajectory is monotonic or follows a sigmodal curve) and aligning subject trajectories to a synchronized timescale based on short-term longitudinal observations (Young et al., 2024). These models are easy to interpret, requiring fewer data points for model fitting. They can be generally categorized into discrete models of biomarker abnormalities and continuous models of biomarker dynamics (see Young et al. (2024) for a more detailed review). Nevertheless, these statistical models often fail to capture the complexity of biomarker trajectories and their dynamic non-linear relationships.

AI techniques, particularly deep learning, have enabled a more efficient data-driven search of biomarkers for the diagnosis and prognosis of AD (Li et al., 2021). Existing multi-modal and speech-based deep learning approaches have primarily focused on symptomatic LOAD diagnosis rather than prognosis (De la Fuente Garcia et al., 2020; Khojaste-Sarakhsi et al., 2022). More recently, several data-driven LOAD prognosis studies have been proposed. Most of these studies have focused on predicting conversions or changes in cognitive scores over time using deep neural networks (Al Olaimat et al., 2023; Ghazi et al., 2019; Jung et al., 2021; Maheux et al., 2023; Nguyen et al., 2023; Wang et al., 2022; Xu et al., 2022). Some studies have also investigated time-to-conversion prediction using deep learning models tailored for survival analysis (Mirabnahrazam et al., 2023; Yi et al., 2023). Moreover, unsupervised learning methods have been developed to derive a prognostic index to predict LOAD progression based on low-cost multi-modal data (Burkhart et al., 2024; Lee et al., 2024). However, these data-driven multi-modal models are often constrained by limited data modalities and features, especially overlooking (1) high-dimensional genetic data (often involving millions of genetic mutation biomarkers) that can improve the biological understanding of LOAD progression and (2) connected speech data that can enable low-cost, non-invasive, and large-scale deployment for LOAD prognosis (De la Fuente Garcia et al., 2020; Elazab et al., 2024; Mueller et al., 2018). They have yet to fully unlock the potential of deep learning in (1) fusing high-dimensional multi-modal data and capturing their complex non-linear relationships for LOAD prognosis, and (2) processing connected speech data for lightweight CL-based LOAD prognosis using simple multi-modal data.

Furthermore, medical AI models are generally data-intensive, often suffering from small, fragmented, heterogeneous, or missing data points (Elazab et al., 2024). To work with limited data, existing deep learning methods for LOAD progression modeling have used simple imputation techniques (e.g., mean values) (Maheux et al., 2023), incorporated built-in imputation mechanisms during model training (Jung et al., 2021), or utilized self-supervised learning (such as input data reconstruction) against model overfitting due to limited data (Wang et al., 2022). However, addressing the data scarcity problem in longitudinal clinical settings remains challenging. For example, the Alzheimer’s Disease Neuroimaging Initiative (ADNI) database is a comprehensive longitudinal dataset, including a wide range of data modalities, such as whole-genome sequencing (WGS), demographics, cognitive assessments, biospecimen, and brain imaging, covering more than 800 subjects who were followed up in multiple years. Many longitudinal features, such as cognitive scores, are sparse and incomplete (Aghili et al., 2022), and some modalities are completely unavailable, especially connected speech (Mueller et al., 2018), which has been utilized to develop low-cost and non-invasive linguistic biomarkers for early detection of AD (Eyigoz et al., 2020). This data scarcity problem has made it difficult for medical AI models to characterize individual disease progression and biomarker changes for accurate prognosis of LOAD.

Recent advancements in generative AI (GAI) and large language models (LLMs), such as ChatGPT, have offered new perspectives to tackle this data scarcity challenge. General-purpose LLMs pre-trained on massive amounts of data from the internet can encode biomedical knowledge and pass medical exams (Singhal et al., 2023). Some also allow for multi-modal inputs, such as electronic health records (EHRs) and imaging (Moor et al., 2023; Thirunavukarasu et al., 2023). One key feature of GAI is generating new data samples that are synthetic yet realistic, making it promising to tackle the data scarcity challenge (Bansal et al., 2022). Some studies have investigated using LLMs for data generation in low-data scenarios, leveraging the prior knowledge of LLMs pre-trained on a large amount of data across different domains (Borisov et al., 2022; Seedat et al., 2023). Given that data generation may also lead to noisy samples, the reliability and utility of newly generated data have also been investigated to assess if they benefit the downstream prediction tasks (Seedat et al., 2023). These studies have provided new insights into building more accurate and reliable AI models in low-data settings with the help of GAI.

This study proposes an AI-driven longitudinal multi-modal platform with time-series transformer (LMP-TX) to overcome the multi-modal data fusion and data scarcity challenges for the early prognosis of LOAD. It has two versions: LMP-TX utilizes full multi-modal data to provide more accurate prediction, while a lightweight version, LMP-TX-CL, only uses simple multi-modal and cognitive-linguistic (CL) data. The key novelties of this work are as follows:

- We exploit LLM-based data generation and imputation techniques to fill in missing multi-modal longitudinal data, including key cognitive and CL data.
- We develop a domain-specific deep neural network model to pre-select the most salient genetic biomarkers indicative of early-stage LOAD progression timings.
- We develop a multi-modal time-series transformer that fuses different modalities and features to predict the timings of early-stage LOAD onsets and identify crucial biomarkers driving early-stage LOAD progressions.

The rest of this paper is organized as follows. Section 2 details the data used in this study and the proposed methodology. Section 3 lists the experimental settings and presents the results. Section 4 discusses the key findings and future work. Section 5 concludes this study.

## 2. Data and Methodology

This study proposes LMP-TX for the early prognosis of LOAD. Specifically, it aims to predict the timings of early-stage LOAD onsets and identify crucial biomarkers that drive early-stage LOAD progressions. The proposed methodology has five steps. First, a comprehensive multi-modal LOAD dataset (but without CL data) was obtained from the ADNI database. In parallel, a simple multi-modal LOAD dataset with connected speech samples was obtained from the DementiaBank database to complement the ADNI database. Progressive ADNI subjects with AD endpoints were selected, and multi-modal data were preprocessed. Second, using LLM-based prompting techniques, CL markers were extracted from the DementiaBank database and used to impute the missing CL data in the ADNI database. Other missing values (e.g., cognitive scores) in the ADNI database were also imputed using LLM-based data generation. Third, the top genetic biomarkers were selected from the ADNI high-dimensional genetic data. Fourth, the preprocessed and imputed multi-modal data were used to generate a large longitudinal dataset in tabular format. A multi-modal time-series transformer model was developed for LOAD progression timing prediction. Finally, feature importance analysis was performed to identify the key multi-modal predictors indicative of different stages of LOAD progressions. Figure 1 shows an overview of the proposed methodology.

**Figure 1.**
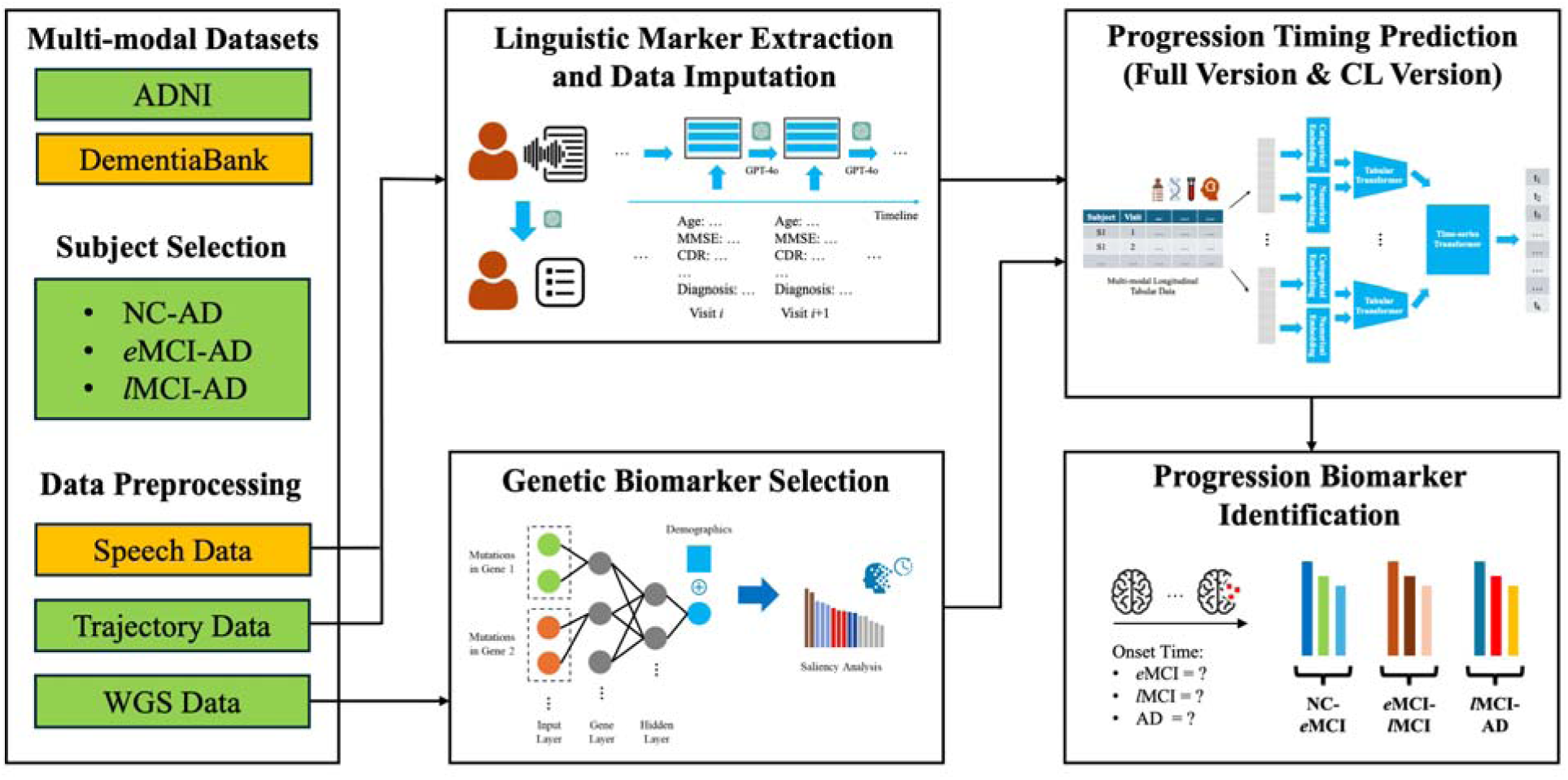
Methodology Overview

### 2.1 Subject Selection and Data Preprocessing

Two datasets were used in this study. The main dataset was obtained from the ADNI database (adni.loni.usc.edu). In the ADNI study, subjects were examined at the baseline visit and tracked via multiple follow-up visits. They were labeled normal control (NC), early mild cognitive impairment (*e*MCI), late mild cognitive impairment (*l*MCI), and AD at each visit. In particular, the *e*MCI and *l*MCI labels were defined by ADNI to improve MCI staging (Edmonds et al., 2019). 185 Caucasian and non-Hispanic/Latino progressive subjects with AD endpoints were selected. Based on their diagnosis at the baseline visit, they were categorized into three groups: NC-AD (*n*=24), *e*MCI-AD (*n*=32), and *l*LMCI-AD (*n*=129). Each subject’s clinical states were assumed to follow the same trajectory, starting from NC and progressing to *e*MCI, *l*MCI, and AD sequentially. Based on the diagnosis labels and dates at each clinical visit, onset timings were calculated for each subject (see Table 1 for the summary statistics). The maximum onset time for *e*MCI, *l*MCI, and AD were 10, 12, and 13 years, respectively.

**Table 1.**
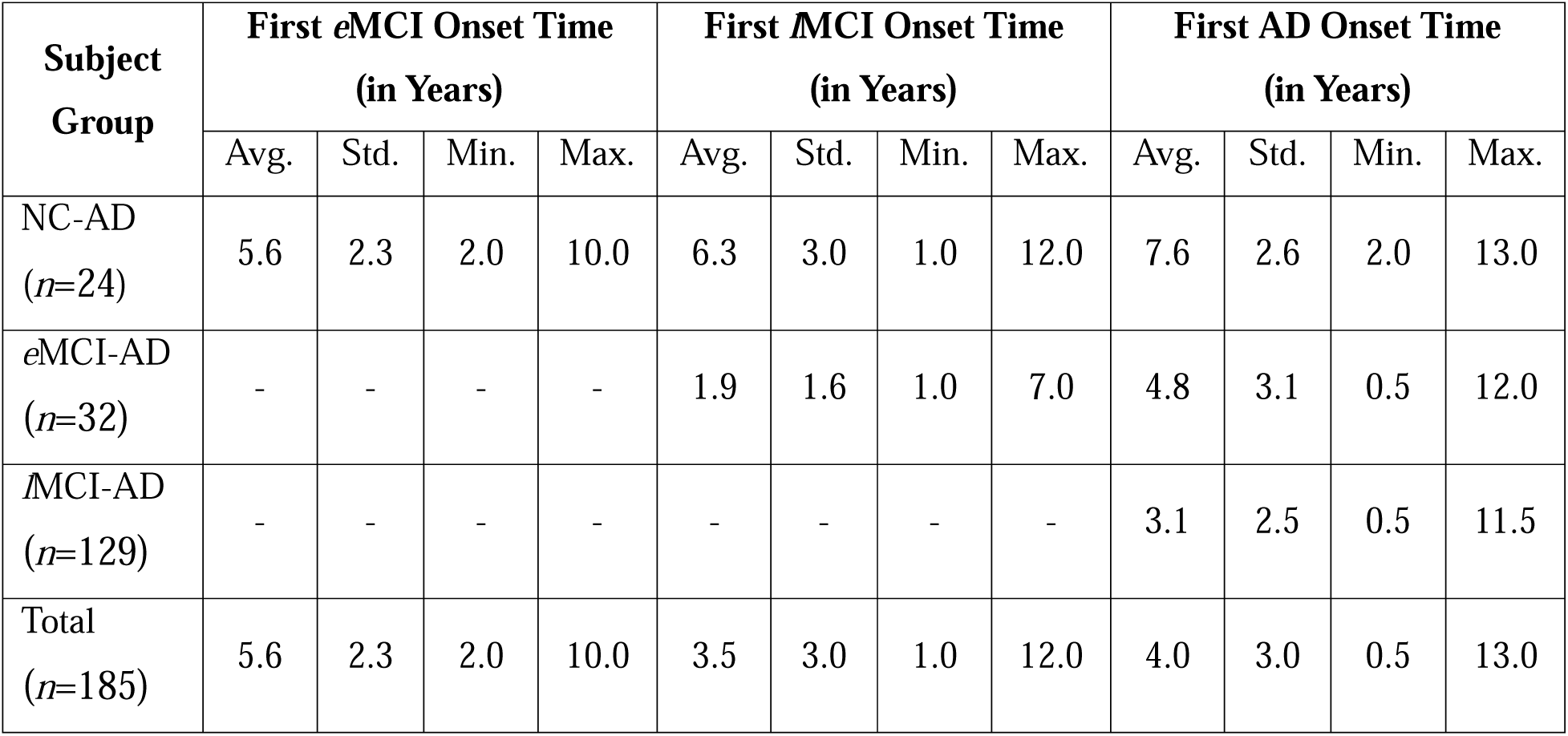
Onset Timing Statistics.

Multi-modal longitudinal (trajectory) data were retrieved from the ADNI database. The following non-genetic data were obtained: (1) demographics and comorbidities (age, gender, family history, and comorbidities), (2) key cognitive assessments, including Mini Mental State Examination (MMSE), Clinical Dementia Rating (CDR), Montreal Cognitive Assessment (MoCA), Alzheimer’s Disease Assessment Scale – Cognitive subscale (ADAS-Cog), and specific items in cognitive assessments that are related to language function (e.g., fluency in MoCA), (3) blood and cerebrospinal fluid (CSF) biomarkers, including ABeta40, ABeta42, P-Tau181, neurofilament light (NFL), proteomics, and metabolomics, (4) imaging, including MRI volumes, thicknesses, and surface areas, PET Amyloid standardized uptake value ratios (SUVRs), and PET Tau SUVRs. 2,110 multi-modal non-genetic features were selected. LLM-based data imputation was performed to impute missing values in the ADNI database (see Section 2.2 for more details).

Moreover, whole-genome sequencing (WGS) data from the ADNI database were preprocessed using standard genetic quality control and linkage disequilibrium (LD)-pruning methods. Genetic variant data from germline and somatic callers were used to incorporate both germline and somatic mutation biomarkers for AD (Downey et al., 2022). 1,229,413 germline variants and 1,495,732 somatic variants were kept after preprocessing. Deep learning-based variant selection was performed to select the top genetic biomarkers (see Section 2.3 for more details).

The second dataset was obtained from the DementiaBank database (dementia.talkbank.org). The DementiaBank database is a speech-based dementia study (Lanzi et al., 2023) that can complement the ADNI database. Connected speech audio samples from the DementiaBank Pitt English corpus (Becker et al., 1994) were converted into transcripts using OpenAI’s Whisper model (OpenAI, n.d.-a). Speaker diarization (Bredin, 2023) was used to remove the examiner’s speech. Each audio sample was associated with the corresponding subject information, including age, gender, MMSE, CDR, and diagnosis label (NC/MCI/AD). LLM-based CL marker extraction was performed to impute missing CL data in the ADNI database (see Section 2.2 for more details).

### 2.2 LLM-based CL Marker Extraction and Longitudinal Data Imputation

Many longitudinal observations, such as cognitive scores, are sparse and incomplete in the ADNI database, and connected speech data are completely unavailable. LLM-based prompting techniques (Borisov et al., 2022; Seedat et al., 2023) were utilized to impute CL and other missing data, exploiting the prior knowledge encoded in LLMs pre-trained on large amounts of data to tackle this data scarcity challenge. OpenAI’s GPT-4o model was used (OpenAI, n.d.-b). The details are described as follows.

For CL data completely unavailable in the ADNI database, CL markers were first extracted from connected speech (Cookie Theft picture description) data from the DementiaBank database (Lanzi et al., 2023). The extracted CL markers were used to recover missing CL data in the ADNI database. Specifically, based on the connected speech transcripts, LLM was prompted to extract CL markers (linguistic patterns) (Mo et al., 2024), for example, “repetitive descriptions” and “fragmented sentence structure”. The prompting included four parts: (1) background information to provide the AD context, such as variable definitions and ranges; (2) the subject’s connected speech transcript; (3) the connected speech transcripts from subjects of different diagnosis labels to help LLM better understand the distinctive CL markers; and (4) step-by-step instructions to elicit the LLM’s reasoning capability for CL marker extraction. The detailed prompting is listed in Section 1 in the Appendix. The average number of extracted CL markers per DementiaBank Pitt subject/visit was five. Some of these extracted CL markers were of high similarity, e.g., “repeated phrases for emphasis or clarity” and “repetition for emphasis or clarity”. *K*-means clustering was further used to group similar CL markers into the same cluster based on their text embeddings, and the optimal number of clusters (*K*=96) was determined using the Silhouette coefficient (Rousseeuw, 1987). Finally, for each ADNI subject at each visit, CL markers associated with similar characteristics (i.e., age, gender, education, MMSE, CDR, and diagnosis label, the common information between ADNI and DementiaBank Pitt) were matched for longitudinal CL data imputation. Based on the observed frequencies of the matched CL markers, the top five CL markers were selected for each ADNI subject/visit using a frequency-weighted randomized sampling approach.

For missing data in the ADNI database, LLM-based longitudinal data imputation was performed recurrently (see Figure 2). Specifically, LLM was prompted to predict one subject’s characteristics during the current visit, and the predicted values were used to fill in missing values and fed into the LLM for the next visit prediction. The prompting included three parts: (1) background information to provide the AD context, such as the variable definitions and ranges; (2) subject information, including demographic information and longitudinal clinical records; and (3) step-by-step instructions to elicit the LLM’s reasoning capability for missing value recovery. The detailed prompting is listed in Section 2 in the Appendix. The newly generated data were used to fill in missing values. Key cognitive variables in the ADNI tabular dataset, including MMSE, MoCA, CDR, and ADAS-Cog, were imputed.

**Figure 2.**
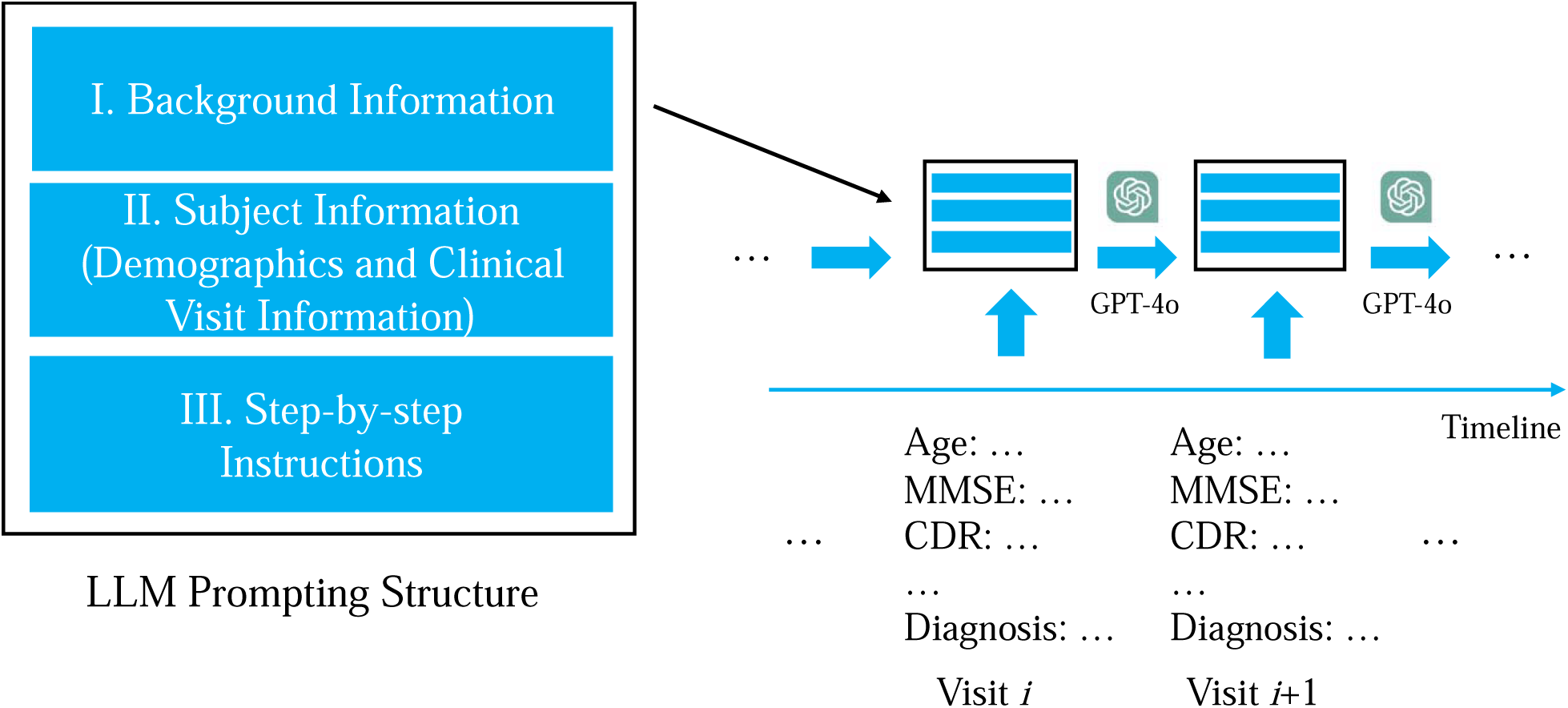
LLM-based Longitudinal Data Imputation

### 2.3 Genetic Biomarker Selection from High-dimensional WGS Data

Preprocessed WGS data still had large amounts of genetic biomarkers (i.e., millions of variants or mutations), making it computationally challenging to incorporate them into LMP-TX. A domain-specific deep feedforward neural network model was developed to pre-select the most salient genetic biomarkers indicative of early-stage LOAD progression timings (see Figure 3). Specifically, each subject was represented by a large high-dimensional feature vector (consisting of millions of features) at the input layer. Each element in the vector indicated whether the subject carried a particular genetic variant (mutation). WGS variants were first clustered by genes. These variants were then connected to the corresponding neurons in the first hidden layer, each representing one gene. This hierarchical design can utilize the sparse connections between the input layer and the first hidden layer, mimicking the gene-based WGS analysis while significantly reducing the number of parameters compared to fully connected layers (Kassani et al., 2022; Zhang et al., 2023). The later hidden layers were standard fully connected layers using the ReLU activation function to learn complex non-linear relationships from the grouped genetic data. The output, a high-level genetic representation vector, was concatenated to the subject’s demographic information (age and gender) for the final prediction. A binary classification was performed based on the concatenated vector to predict whether the subject was a slow or fast progressor using a two-year cut-off of the corresponding onset timing. The onset event was *e*MCI and *l*MCI for NC and *e*MCI subjects, respectively. Feature importance analysis wa performed to identify the most important genetic biomarkers predicting slow or fast progression using the saliency scores (Simonyan, 2013) calculated based on the trained genetic model. The top *K* (*K*=100) genetic biomarkers were selected.

**Figure 3.**
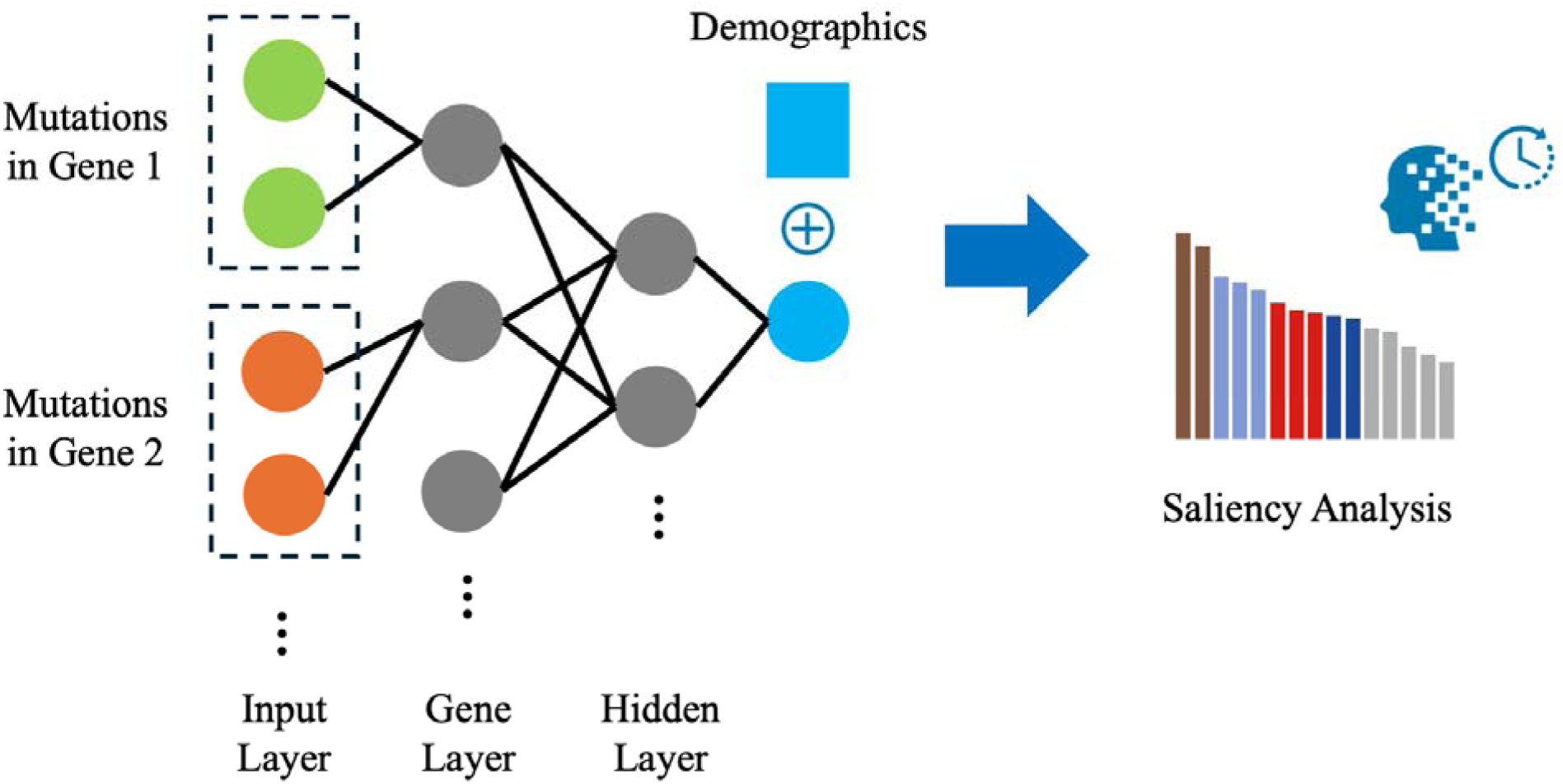
Domain-Specific Deep Neural Network for Genetic Biomarker Selection

### 2.4 LOAD Progression Timing Prediction

The preprocessed subject trajectory data were combined with the top genetic biomarker (unchanged over multiple visits) to generate a large longitudinal dataset in tabular format. Each row included one subject’s information recorded at one clinical visit, including multi-modal covariates and outcomes (onset timings). Two versions of LMP-TX were developed using the same methodology. The full version utilized all available multi-modal data (demographics, CL biomarkers, genetic biomarkers, cognitive tests, blood/CSF biomarkers, and imaging biomarkers). The CL version only used CL data plus simple multi-modal data (e.g., demographics, MMSE, MoCA, and blood biomarkers). The detailed methodology is elaborated as follows (see Figure 4).

**Figure 4.**
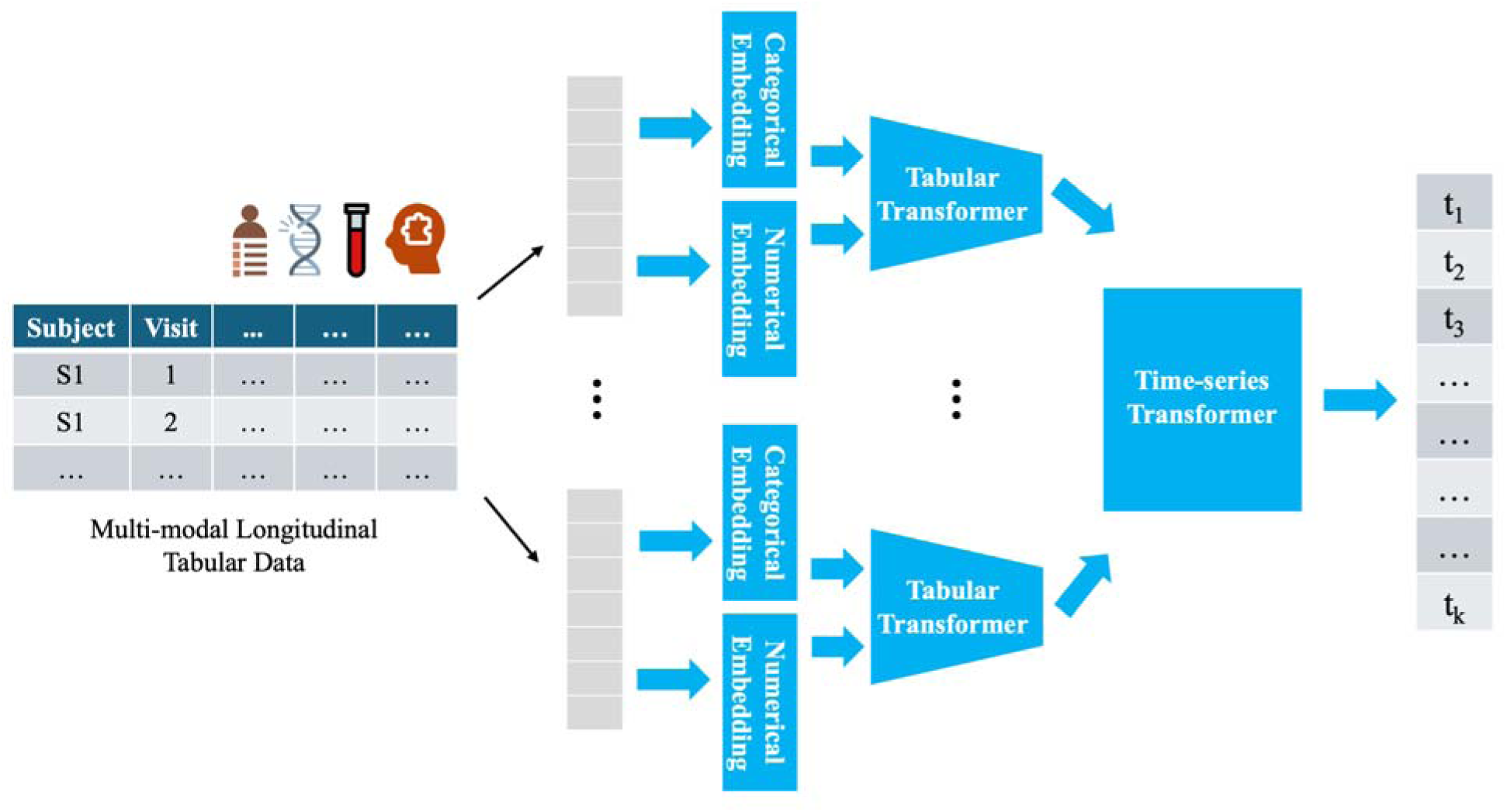
Multi-modal Time-series Transformer for LOAD Progression Timing Prediction

A time-series transformer-based framework was developed for progression timing prediction. Specifically, a transformer-based encoder (Gorishniy et al., 2021) was adopted to embed multi-modal tabular data into the same latent space while capturing their shared and specific semantics. Categorical and numerical values were first mapped to embeddings as the transformer model inputs. A self-supervised learning procedure was adopted to predict one masked part of the input based on the remaining input, making the encoder capable of addressing noisy/missing data by design and capturing the contextual representations of multi-modal features. After the pre-training process, the pre-trained encoder generated a high-level multi-modal representation for each subject at each visit. The representations obtained from the current and previous visits were fed into a time-series transformer for longitudinal data modeling. Fully connected layers were used to predict the final outcomes, i.e., the probability of onset time (from 1 to *T*, where *T* was determined by the maximum period observed in the dataset). Onset time was represented by disjoint time windows, , , , …, , to model the onset risk over time. A binary classification was performed to predict the onset event in a given *k*-year time window (Zhang et al., 2020). Three transformer models were trained for each onset event (i.e., *e*MCI, *l*MCI, and AD).

### 2.5 LOAD Progression Biomarker Identification

Feature importance analysis was performed to identify the multi-modal predictors indicative of different stages of LOAD progressions, especially early-stage progressions, including NC-*e*MCI and *e*MCI-*l*MCI. Based on the trained models, Shapley values were computed using the Kernel SHapley Additive exPlanations (SHAP) method (Lundberg, 2017). The average Shapley values across all time windows were used to assess the contributions of different multi-modal features in progression timing prediction.

## 3. Results

### 3.1 Experimental Settings

A 70/30 stratified split of all selected subjects was used for model development and testing, preserving a balanced distribution of NC-AD, *e*MCI-AD, and *l*MCI-AD subjects. The progression prediction model was trained using the following hyper-parameters: batch size (32), learning rate (1e^-3^), and maximum epochs (100). The following hyper-parameters were fine-tuned and selected using a subset of the training data as a validation set: the hidden dimension size (16, 32, or 64), the depth (1 to 4), and the number of multi-head attention (1 to 4). An early stop was adopted if the validation performance was not improved over ten epochs.

### 3.2 Predictive Performance Evaluation

The area under the ROC curve (AUC) score was used to evaluate progression timing predictions at different years (from 1-year prediction up to 10-year prediction). Two versions of LMP-TX were assessed. The full version utilized all multi-modal data, while the CL version utilized only simple multi-modal data. Prognosis accuracy based on the AUC scores for subjects who progressed from NC to *e*MCI and *e*MCI to *l*MCI is respectively 89% maximum (predicted by LMP-TX) and 81% maximum (predicted by LMP-TX-CL) (see Table 2 for more details). In general, predicting *e*MCI onset events is more challenging compared to other LOAD states. When comparing the full version to the CL version, higher predictive accuracy is achieved by including more multi-modal features.

**Table 2.**
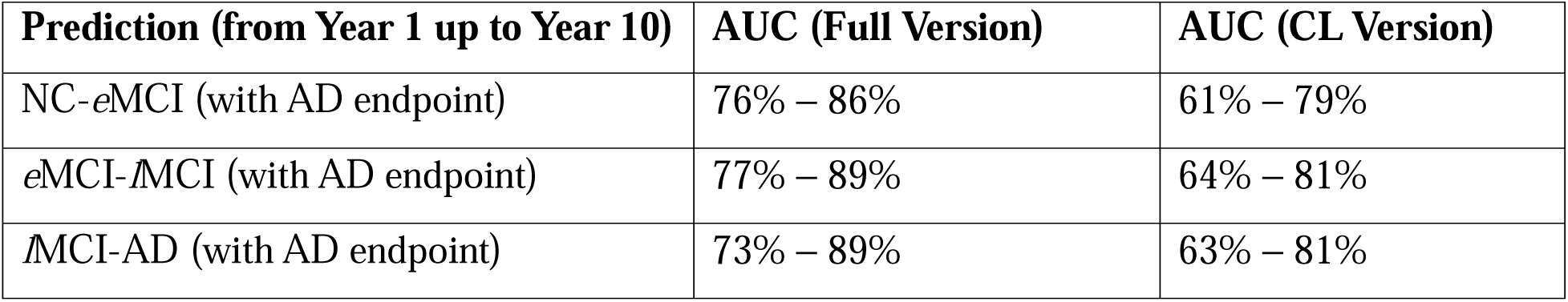
Prognosis Performance.

| Prediction (from Year 1 up to Year 10) | AUC (Full Version) | AUC (CL Version) |
| --- | --- | --- |
| NC-*e*MCI (with AD endpoint) | 76% – 86% | 61% – 79% |
| *e*MCI-*l*MCI (with AD endpoint) | 77% – 89% | 64% – 81% |
| *l*MCI-AD (with AD endpoint) | 73% – 89% | 63% – 81% |

### 3.3 Key Multi-modal Biomarkers Predicting Progression Timings

Tables 3-5 list the results of the top multi-modal biomarkers predicting progression timings at different stages. Among the identified top biomarkers, gender is consistently the most salient at each stage. Triglycerides, a serum biomarker related to lipid metabolism, is also implicated in different stages. The other biomarkers vary across different stages. Specifically, for the NC-*e*MCI stage (see Table 3), the top biomarkers cover various aspects, including genetics, imaging (MRI and PET), biospecimen (proteins and metabolites), and connected speech. The genetic biomarkers, including CBX2 and ST8SIA2, are related to neurodevelopment and have been reported by previous literature on AD and neurodegeneration (Gu et al., 2018; Stefano et al., 2016). The CL-based biomarker (“fragmented sentence structure”) is related to syntactic impairment in AD (Fraser et al., 2015; Lofgren & Hinzen, 2022). Moreover, imaging biomarkers highlight the brain regions implicated in the early AD stage, including the superior frontal cortex (Keith et al., 2023), frontal pole cortex (Finger et al., 2017), and left superior parietal (Hänggi et al., 2011). Further, the identified serum metabolite biomarkers, including very-low-density lipoprotein and triglycerides, are related to brain inflammation and metabolism (Bernath et al., 2020; Lin et al., 2021), while the CSF protein biomarker, CO8B, is involved in the innate immunity complement system and related to inflammatory responses in AD (Fukuda et al., 2024).

**Table 3.** Top Biomarkers Predicting NC-*e*MCI Progression.

| Rank | Modality | Feature |
| --- | --- | --- |
| 1 | Demographics | Gender (male) |
| 2 | Connected Speech | CL Marker (fragmented sentence structure) |
| 3 | Genetics | Gene (CBX2) |
| 4 | Imaging | PET Tau SUVR (superior frontal cortex) |
| 5 | Imaging | MRI (cortical volume of left superior parietal) |
| 6 | Biospecimen | CSF Protein (CO8B) |
| 7 | Biospecimen | Serum Metabolite (very-low-density lipoprotein) |
| 8 | Genetics | Gene (ST8SIA2) |
| 9 | Imaging | PET Amyloid SUVR (frontal pole cortex) |
| 10 | Biospecimen | Serum Metabolite (triglycerides) |

For the *e*MCI-*l*MCI stage (see Table 4), the top genetic biomarkers are mostly related to the immune system and neuroinflammation: LINC01162 is a long non-coding RNA (lncRNA) gene involved in B-cell differentiation (Lagou et al., 2018) and ATP2A2 is related to calcium homeostasis (Lim et al., 2021) which is linked to neuroinflammation (Sama & Norris, 2013). The imaging biomarkers reveal the left lingual region related to visual processing and faster cognitive decline (Meulenbroek et al., 2010) and the left pallidum region implicated in brain network topology changes in late MCI converters (Pereira et al., 2016). Further, the serum metabolite biomarkers include serum short-chain fatty acids (hexanoic acid and valeric acid) that can mediate gut-brain interactions and affect neuroinflammation (Qian et al., 2022), serum phosphatidylcholine related to choline metabolism (Whiley et al., 2014), and CSF protein CRP related to inflammation (Brosseron et al., 2018).

**Table 4.** Top Biomarkers Predicting *e*MCI-*l*MCI Progression.

| Rank | Modality | Feature |
| --- | --- | --- |
| 1 | Demographics | Gender (male) |
| 2 | Biospecimen | CSF Protein (CRP) |
| 3 | Genetics | Gene (LINC01162) |
| 4 | Biospecimen | Serum Metabolite (hexanoic acid) |
| 5 | Biospecimen | Serum Metabolite (valeric acid) |
| 6 | Imaging | MRI (surface area of left lingual) |
| 7 | Biospecimen | Serum Metabolite (triglycerides) |
| 8 | Imaging | PET Tau SUVR (left pallidum) |
| 9 | Genetics | Gene (ATP2A2) |
| 10 | Biospecimen | Serum Metabolite (phosphatidylcholine) |

**Table 5.** Top Biomarkers Predicting *l*MCI-AD Progression.

| Rank | Modality | Feature |
| --- | --- | --- |
| 1 | Demographics | Gender (male) |
| 2 | Imaging | PET Amyloid SUVR (brain stem) |
| 3 | Biospecimen | Serum Metabolite (palmitoleic acid) |
| 4 | Biospecimen | Serum Metabolite (docosatetraenoic acid) |
| 5 | Genetics | Gene (BANK1) |
| 6 | Imaging | MRI (cortical thickness of right rostral anterior cingulate) |
| 7 | Biospecimen | CSF Protein (LYSC) |
| 8 | Biospecimen | Serum Metabolite (triglycerides) |
| 9 | Biospecimen | CSF Protein (PCSK1) |
| 10 | Biospecimen | CSF Protein (RET4) |

For the *l*MCI-AD stage (see Table 5), the top genetic biomarker, BANK1, encodes a B-cell-specific protein related to the immune system (Blokland et al., 2017). Imaging biomarkers include the brain stem and the right rostral anterior cingulate regions related to cognitive aging (Haller et al., 2020; Pezzoli et al., 2024). Serum metabolites include fatty acids (palmitoleic acid and docosatetraenoic acid) that have exhibited dysregulation in AD cases (Hosseini et al., 2020). CSF proteins include LYSC, PCSK1, and RET4. Some of them (e.g., PCSK1) have been found to improve AD diagnosis and prognosis accuracy (Guo et al., 2024).

## 4. Discussion and Future Work

This study proposes LMP-TX for early LOAD prognosis. LMP-TX uses LLM-based data imputation techniques and a transformer model for longitudinal multi-modal data fusion. Two versions of LMP-TX have been developed, including a full version using all available multi-modal data and a lightweight version, LMP-TX-CL, which utilizes simple multi-modal and CL data. Results have demonstrated the effectiveness of LMP-TX in providing accurate predictions for LOAD progressions at different stages. Prognosis accuracy based on the AUC scores for subjects progressing from NC to *e*MCI and *e*MCI to *l*MCI is respectively 89% maximum (predicted by LMP-TX) and 81% maximum (predicted by LMP-TX-CL).

Moreover, results have revealed key multi-modal biomarkers indicative of early-stage LOAD progressions. These biomarkers, covering genetics, imaging, metabolites, proteomics, and connected speech, highlight the diversity of the biological processes involved in LOAD and the complex interplay of biomarkers across different stages of LOAD. An interesting finding about the connected speech-based CL marker is the identification of fragmented sentence structure as a biomarker for early-stage LOAD progression. This linguistic feature can be related to syntactic impairment in LOAD (Fraser et al., 2015; Lofgren & Hinzen, 2022), which can be used as an early, non-invasive marker of cognitive decline, offering a novel way to assess AD-related neurodegeneration through natural language processing. This finding highlights the potential of linguistic-based biomarkers in detecting subtle cognitive changes even before more pronounced memory deficits appear. Moreover, early-stage biomarkers highlight neurodevelopmental genes and dysregulation in metabolic and inflammatory pathways. As the disease progresses, neuroinflammation, immune responses, and metabolic dysfunction become more pronounced. These multi-modal biomarkers provide crucial insights into the pathophysiology of LOAD, opening new avenues for early prognosis and targeted therapeutic interventions.

However, this study has some limitations that can be addressed in future work. First, this study mainly focuses on the ADNI dataset. However, AI models require large datasets integrating genetics, imaging, biospecimen, and connected speech data to achieve high generalizability. Such comprehensive datasets are rare, especially those with speech modality covering different stages of LOAD progression. The lack of high-quality multi-modal datasets limits the generalizability of AI models. Future work will incorporate more LOAD datasets, such as those from the Alzheimer’s Disease Sequencing Project – Phenotype Harmonization Consortium (ADSP-PHC) and from UK Biobank, to improve and validate the generalizability of the proposed AI approach. Second, this study is based on data from a specific population (i.e., Caucasian), which may not represent the full spectrum of AD across different ethnicities and genetic backgrounds. To date, the study subjects in AD datasets have been mostly Caucasians, and other ethnicities are often underrepresented. As a result, AI models may not generalize well to disease progressions among the underrepresented, leading to biased prediction. For example, lower sensitivity was observed for predicting MCI to AD conversion among certain minority ethnic groups in the ADNI study (Yuan et al., 2023). In the past, transfer learning techniques have been adopted for cross-population learning. For example, English corpus data has facilitated speech-based AD detection using the Chinese AD corpus data (Guo et al., 2020). Future work will investigate transfer learning techniques to improve cross-population LOAD prognosis limited by low-resource data.

Finally, in future work, a more fine-grained LMP-TX will be developed. Clinical stages, such as *e*MCI, *l*MCI, and AD used in this study, are discrete and less fine-grained labels, often depending on cognitive scores and failing to fully capture the pre-symptomatic phase of LOAD. A Disease Progression Score (DPS), ranging from 0 (corresponding to NC) to 1 (corresponding to AD), will be calculated to re-label each subject’s discrete diagnosis along the AD continuum at each visit based on the corresponding progression timing predictors. Specifically, based on the initial four labels (NC, *e*MCI, *l*MCI, and AD) of the ADNI dataset, the DPS scale will be divided into three intervals, namely, NC-*e*MCI, *e*MCI-*l*MCI, and *l*MCI -AD. The cut-offs to determine the three intervals will be optimized to separate different labels. Given the most salient biomarkers during these three stages shown in Section 3.3, for each interval per subject, the corresponding most salient biomarkers will be used to determine a DPS value. As a result, someone who was labeled NC originally but with higher biomarker values will be placed on the DPS scale closer to *e*MCI than to NC. After DPS calculation, LMP-TX will be re-trained to predict DPS over time. The rate of change in DPS at different stages will be calculated. A pre-symptomatic state between NC and *e*MCI can also be determined based on the maximum rate of change in progression. Finally, feature importance analysis will be performed based on the trained fine-grained LMP-TX model for DPS prediction to determine the most important biomarkers driving fast progressions at early stages. Candidate drugs and combinations targeting early-stage LOAD progression-driven biomarkers will be identified and verified (Li et al., 2024).

## 5. Conclusion

Early prognosis of LOAD is critical for timely intervention. This study proposes LMP-TX, an AI-driven integrated longitudinal multi-modal platform for the early prognosis of LOAD. It exploits LLM-driven data imputation techniques to fill in missing multi-modal data, including CL data. It utilizes a transformer model to fuse different modalities and features from longitudinal multi-modal data to predict the timings of LOAD progressions at early stages and identify the key biomarkers deterministic of early-stage LOAD progression timings. Results have demonstrated up to 89% accuracy for progression timing prediction and revealed key multi-modal biomarkers driving early-stage LOAD progressions. Future work will use the identified biomarkers during different LOAD progression stages, especially the early stages, to calculate disease progression scores, develop a more fine-grained LMP-TX for predicting disease progression scores, and identify the key multi-modal and CL-based biomarkers predictive of fast AD progression rates at early stages.

## Data Availability

The ADNI database is available at https://adni.loni.usc.edu. The DementiaBank database is available at https://dementia.talkbank.org.

## Competing Interests

The authors declare no competing interests.

## Funding Statement

This research was supported in part by the US National Academy of Medicine (NAM) Healthy Longevity Catalyst Award 2023, administered by the Research Grants Council of Hong Kong under Grant No. HLCA/E-712/23, and in part by the HKU Seed Funding for Collaborative Research, June 2023. The funder had no role in study design, data collection and analysis, decision to publish, or preparation of the manuscript.

## Acknowledgements

The authors are grateful for the Pitt corpus provided by the DementiaBank database. The authors would also like to acknowledge the following grant support for the Pitt corpus: NIA AG03705 and AG05133. The authors would also like to acknowledge the following grant support for the ADNI database. Data collection and sharing for the Alzheimer’s Disease Neuroimaging Initiative (ADNI) is funded by the National Institute on Aging (National Institutes of Health Grant U19AG024904). The grantee organization is the Northern California Institute for Research and Education. In the past, ADNI has also received funding from the National Institute of Biomedical Imaging and Bioengineering, the Canadian Institutes of Health Research, and private sector contributions through the Foundation for the National Institutes of Health (FNIH) including generous contributions from the following: AbbVie, Alzheimer’s Association; Alzheimer’s Drug Discovery Foundation; Araclon Biotech; BioClinica, Inc.; Biogen; Bristol-Myers Squibb Company; CereSpir, Inc.; Cogstate; Eisai Inc.; Elan Pharmaceuticals, Inc.; Eli Lilly and Company; EuroImmun; F. Hoffmann-La Roche Ltd and its affiliated company Genentech, Inc.; Fujirebio; GE Healthcare; IXICO Ltd.; Janssen Alzheimer Immunotherapy Research & Development, LLC.; Johnson & Johnson Pharmaceutical Research & Development LLC.; Lumosity; Lundbeck; Merck & Co., Inc.; Meso Scale Diagnostics, LLC.; NeuroRx Research; Neurotrack Technologies; Novartis Pharmaceuticals Corporation; Pfizer Inc.; Piramal Imaging; Servier; Takeda Pharmaceutical Company; and Transition Therapeutics. Data collection and sharing for this project was funded by the Alzheimer’s Disease Metabolomics Consortium (National Institute on Aging R01AG046171, RF1AG051550 and 3U01AG024904-09S4). Data generation and sharing for this project was funded, in part, by the Alzheimer’s Disease Sequencing Project. Data used in preparation of this article were obtained from the Alzheimer’s Disease Neuroimaging Initiative (ADNI) database (adni.loni.usc.edu). As such, the investigators within the ADNI contributed to the design and implementation of ADNI and/or provided data but did not participate in analysis or writing of this report. A complete listing of ADNI investigators can be found at: http://adni.loni.usc.edu/wp-content/uploads/how_to_apply/ADNI_Acknowledgement_List.pdf

## Appendix

### 1. LLM Prompting Example for CL Marker Extraction

*Background information: Three labels are used to indicate one’s diagnosis: Normal Control (NC), Mild Cognitive Impairment (MCI), and Alzheimer’s Disease (AD). Clinical Dementia Rating (CDR) ranges from 0 to 5. Mini-Mental State Examination (MMSE) ranges from 0 to 30. Missing values are denoted as nan*.

*One subject is diagnosed as MCI at Age: [age]. This subject’s demographic information:*

*Gender: [gender], Years of Education: [education]*

*MMSE: [MMSE], CDR: [CDR]*

*This subject’s connected speech transcript: There’s a young boy that’s getting a cookie jar …*

*Some examples extracted from other subjects:*

*NC subject connected speech transcript example 1: Well, the mother’s doing the dishes …*

*NC subject connected speech transcript example 2: …*

*NC subject connected speech transcript example 3: …*

*AD subject connected speech transcript example 1: The sink is running over …*

*AD subject connected speech transcript example 2: …*

*AD subject connected speech transcript example 3: …*

*Identify the linguistic markers that could potentially indicate MCI at Age: [age], based on the information above. The linguistic markers should be extracted from the subject’s connected speech transcript provided above. The identified linguistic markers should not overlap with those extracted from the examples of NC and AD subjects above*.

*Output the identified linguistic markers in JSON format as follows*.

*Firstly, extract a list of linguistic patterns that characterize the subject’s cognitive health status in the field named PATTERNS*.

*Secondly, extract a list of linguistic keywords or phrases that characterize the subject’s cognitive health status in the field named KEYWORDS/PHRASES*.

*Thirdly, provide a brief explanation of the linguistic markers extracted in the field named EXPLANATION*.

*A JSON schema is provided as follows:*

~~~
*{
 “PATTERNS“: [
  “string“
 ],
 “KEYWORDS/PHRASES“: [
  “string“
 ],
 “EXPLANATION“:
  “string“
}*
~~~

### 2. LLM Prompting Example for Longitudinal Data Imputation

*Background information: Three labels are used to indicate one’s diagnosis: Normal Control (NC), Mild Cognitive Impairment (MCI), and Alzheimer’s Disease (AD). Clinical Dementia Rating (CDR) ranges from 0 to 5. Mini-Mental State Examination (MMSE) ranges from 0 to 30. Montreal Cognitive Assessment (MoCA) ranges from 0 to 30. Alzheimer’s Disease Assessment Scale-Cognitive Subscale (ADAS-Cog) ranges from 0 to 70. Missing values are denoted as nan*.

*One subject is diagnosed as MCI at Age: [age]. This subject’s demographic information:*

*Gender: [gender], Education (in years): [education]*

*This subject’s clinical visit history:*

*Age: […]*

*Mini-Mental State Examination (MMSE): […]*

*Clinical Dementia Rating (CDR): […]*

*Montreal Cognitive Assessment (MoCA): […]*

*Alzheimer’s Disease Assessment Scale - Cognitive Subscale (ADAS-Cog): […]*

*Diagnosis Label: […]*

*Predict this subject’s missing values at Age: [age], based on the information above*.

*Do it step by step and output in JSON format as follows*.

*Predict the subject’s diagnosis using ’NC’, ’MCI’, or ’AD’ in the field named LABEL*.

*Predict the subject’s MMSE score in the field named MMSE*.

*Predict the subject’s CDR score in the field named CDR*.

*Predict the subject’s MoCA score in the field named MoCA*.

*Predict the subject’s ADAS-Cog score in the field named ADAS-Cog*.

*Finally, output the reasons for ’LABEL’, ’MMSE’, ’CDR’, ’MoCA’, and ’ADAS-Cog’ in the field named REASONING*.

*A JSON schema is provided as follows:*

~~~
*{
 “LABEL“: “string”,
 “MMSE“: “number”,
 “CDR“: “number”,
 “MoCA“: “number”,
 “ADAS-Cog“: “number”,
 “REASONING“: {
  “LABEL“: “string”,
  “MMSE“: “string”,
  “CDR“: “string”,
  “MoCA“: “string”,
  “ADAS-Cog“: “string“
 }
}*
~~~

## Notes

### Competing Interest Statement

The authors have declared no competing interest.

## References

Aghili, M., Tabarestani, S., & Adjouadi, M. (2022). Addressing the missing data challenge in multi-modal datasets for the diagnosis of Alzheimer’s disease. Journal of Neuroscience Methods, 375, 109582.

Al Olaimat, M., Martinez, J., Saeed, F., Bozdag, S., & Alzheimer’s Disease Neuroimaging Initiative. (2023). PPAD: A deep learning architecture to predict progression of Alzheimer’s disease. Bioinformatics, 39(Supplement_1), i149–i157.

Bansal, M. A., Sharma, D. R., & Kathuria, D. M. (2022). A systematic review on data scarcity problem in deep learning: solution and applications. ACM Computing Surveys, 54(10s), 1–29.

Becker, J. T., Boiler, F., Lopez, O. L., Saxton, J., & McGonigle, K. L. (1994). The natural history of Alzheimer’s disease: description of study cohort and accuracy of diagnosis. Archives of Neurology, 51(6), 585–594.

Bernath, M. M., Bhattacharyya, S., Nho, K., Barupal, D. K., Fiehn, O., Baillie, R., Risacher, S. L., Arnold, M., Jacobson, T., & Trojanowski, J. Q. (2020). Serum triglycerides in Alzheimer disease: Relation to neuroimaging and CSF biomarkers. Neurology, 94(20), e2088–e2098.

Blokland, G. A., Wallace, A. K., Hansell, N. K., Thompson, P. M., Hickie, I. B., Montgomery, G. W., Martin, N. G., McMahon, K. L., de Zubicaray, G. I., & Wright, M. J. (2017). Genome-wide association study of working memory brain activation. International Journal of Psychophysiology, 115, 98–111.

Borisov, V., Seßler, K., Leemann, T., Pawelczyk, M., & Kasneci, G. (2022). Language models are realistic tabular data generators. arXiv preprint arXiv:2210.06280.

Bredin, H. (2023). pyannote. audio 2.1 speaker diarization pipeline: principle, benchmark, and recipe. 24th INTERSPEECH Conference (INTERSPEECH 2023),

Brosseron, F., Traschütz, A., Widmann, C. N., Kummer, M. P., Tacik, P., Santarelli, F., Jessen, F., & Heneka, M. T. (2018). Characterization and clinical use of inflammatory cerebrospinal fluid protein markers in Alzheimer’s disease. Alzheimer’s research & therapy, 10, 1–14.

Burkhart, M. C., Lee, L. Y., Vaghari, D., Toh, A. Q., Chong, E., Chen, C., Tiňo, P., & Kourtzi, Z. (2024). Unsupervised multimodal modeling of cognitive and brain health trajectories for early dementia prediction. Scientific Reports, 14(1), 10755.

De la Fuente Garcia, S., Ritchie, C. W., & Luz, S. (2020). Artificial intelligence, speech, and language processing approaches to monitoring Alzheimer’s disease: a systematic review. Journal of Alzheimer’s Disease, 78(4), 1547–1574.

Downey, J., Lam, J. C. K., Li, V. O. K., & Gozes, I. (2022). Somatic mutations and Alzheimer’s disease. Journal of Alzheimer’s Disease, 90(2), 475–493.

Edmonds, E. C., McDonald, C. R., Marshall, A., Thomas, K. R., Eppig, J., Weigand, A. J., Delano-Wood, L., Galasko, D. R., Salmon, D. P., & Bondi, M. W. (2019). Early versus late MCI: Improved MCI staging using a neuropsychological approach. Alzheimer’s & Dementia, 15(5), 699–708.

Elazab, A., Wang, C., Abdelaziz, M., Zhang, J., Gu, J., Gorriz, J. M., Zhang, Y., & Chang, C. (2024). Alzheimer’s disease diagnosis from single and multimodal data using machine and deep learning models: Achievements and future directions. Expert Systems with Applications, 124780.

Eyigoz, E., Mathur, S., Santamaria, M., Cecchi, G., & Naylor, M. (2020). Linguistic markers predict onset of Alzheimer’s disease. EClinicalMedicine, 28.

Finger, E., Zhang, J., Dickerson, B., Bureau, Y., Masellis, M., & Initiative, A. s. D. N. (2017). Disinhibition in Alzheimer’s disease is associated with reduced right frontal pole cortical thickness. Journal of Alzheimer’s Disease, 60(3), 1161–1170.

Fraser, K. C., Meltzer, J. A., & Rudzicz, F. (2015). Linguistic features identify Alzheimer’s disease in narrative speech. Journal of Alzheimer’s Disease, 49(2), 407–422.

Fukuda, M., Okanishi, H., Ino, D., Ono, K., Kawamura, S., Wakai, E., Miyoshi, T., Sato, T., Ohta, Y., & Saito, T. (2024). Disturbance in the protein landscape of cochlear perilymph in an Alzheimer’s disease mouse model. Plos One, 19(5), e0303375.

Ghazi, M. M., Nielsen, M., Pai, A., Cardoso, M. J., Modat, M., Ourselin, S., Sørensen, L., & Alzheimer’s Disease Neuroimaging Initiative. (2019). Training recurrent neural networks robust to incomplete data: Application to Alzheimer’s disease progression modeling. Medical Image Analysis, 53, 39–46.

Gorishniy, Y., Rubachev, I., Khrulkov, V., & Babenko, A. (2021). Revisiting deep learning models for tabular data. Advances in Neural Information Processing Systems, 34, 18932–18943.

Gu, X., Wang, X., Su, D., Su, X., Lin, L., Li, S., Wu, Q., Liu, S., Zhang, P., & Zhu, X. (2018). CBX2 inhibits neurite development by regulating neuron-specific genes expression. Frontiers in Molecular Neuroscience, 11, 46.

Guo, Y., Chen, S.-D., You, J., Huang, S.-Y., Chen, Y.-L., Zhang, Y., Wang, L.-B., He, X.-Y., Deng, Y.-T., & Zhang, Y.-R. (2024). Multiplex cerebrospinal fluid proteomics identifies biomarkers for diagnosis and prediction of Alzheimer’s disease. Nature Human Behaviour, 1–20.

Guo, Z., Liu, Z., Ling, Z., Wang, S., Jin, L., & Li, Y. (2020). Text classification by contrastive learning and cross-lingual data augmentation for alzheimer’s disease detection. Proceedings of the 28th International Conference on Computational Linguistics,

Haller, S., Montandon, M.-L., Lilja, J., Rodriguez, C., Garibotto, V., Herrmann, F. R., & Giannakopoulos, P. (2020). PET amyloid in normal aging: direct comparison of visual and automatic processing methods. Scientific Reports, 10(1), 16665.

Hänggi, J., Streffer, J., Jäncke, L., & Hock, C. (2011). Volumes of lateral temporal and parietal structures distinguish between healthy aging, mild cognitive impairment, and Alzheimer’s disease. Journal of Alzheimer’s Disease, 26(4), 719–734.

Hosseini, M., Poljak, A., Braidy, N., Crawford, J., & Sachdev, P. (2020). Blood fatty acids in Alzheimer’s disease and mild cognitive impairment: A meta-analysis and systematic review. Ageing Research Reviews, 60, 101043.

Jung, W., Jun, E., Suk, H.-I., & Alzheimer’s Disease Neuroimaging Initiative. (2021). Deep recurrent model for individualized prediction of Alzheimer’s disease progression. NeuroImage, 237, 118143.

Kassani, P. H., Lu, F., Le Guen, Y., Belloy, M. E., & He, Z. (2022). Deep neural networks with controlled variable selection for the identification of putative causal genetic variants. Nature Machine Intelligence, 4(9), 761–771.

Keith, C. M., Haut, M. W., Wilhelmsen, K., Mehta, R. I., Miller, M., Navia, R. O., Ward, M., Lindberg, K., Coleman, M., & McCuddy, W. T. (2023). Frontal and temporal lobe correlates of verbal learning and memory in aMCI and suspected Alzheimer’s disease dementia. *Aging*, Neuropsychology, and Cognition, 30(6), 923–939.

Khojaste-Sarakhsi, M., Haghighi, S. S., Ghomi, S. F., & Marchiori, E. (2022). Deep learning for Alzheimer’s disease diagnosis: A survey. Artificial Intelligence in Medicine, 130, 102332.

Lagou, V., Garcia-Perez, J. E., Smets, I., Van Horebeek, L., Vandebergh, M., Chen, L., Mallants, K., Prezzemolo, T., Hilven, K., & Humblet-Baron, S. (2018). Genetic architecture of adaptive immune system identifies key immune regulators. Cell Reports, 25(3), 798–810. e796.

Lanzi, A. M., Saylor, A. K., Fromm, D., Liu, H., MacWhinney, B., & Cohen, M. L. (2023). DementiaBank: Theoretical rationale, protocol, and illustrative analyses. American Journal of Speech-Language Pathology, 32(2), 426–438.

Lee, L. Y., Vaghari, D., Burkhart, M. C., Tino, P., Montagnese, M., Li, Z., Zühlsdorff, K., Giorgio, J., Williams, G., Chong, E., Chen, C., Underwood, B. R., Rittman, T., & Kourtzi, Z. (2024). Robust and interpretable AI-guided marker for early dementia prediction in real-world clinical settings. EClinicalMedicine, 74. 10.1016/j.eclinm.2024.102725

Li, V. O. K., Han, Y., Kaistha, T., Zhang, Q., Downey, J., Gozes, I., & Lam, J. C. K. (2024). DeepDrug: An Expert-led Domain-specific AI-Driven Drug-Repurposing Mechanism for Selecting the Lead Combination of Drugs for Alzheimer’s Disease. medRxiv, 2024.2007. 2006.24309990.

Li, V. O. K., Lam, J. C. K., Han, Y., Cheung, L. Y. L., Downey, J., Kaistha, T., & Gozes, I. (2021). Designing a protocol adopting an artificial intelligence (AI)–driven approach for early diagnosis of late-onset Alzheimer’s disease. Journal of Molecular Neuroscience, 71(7), 1329–1337.

Lim, B., Prassas, I., & Diamandis, E. P. (2021). Alzheimer disease pathogenesis: the role of autoimmunity. The Journal of Applied Laboratory Medicine, 6(3), 756–764.

Lin, Y.-S., Liu, C.-K., Lee, H.-C., Chou, M.-C., Ke, L.-Y., Chen, C.-H., & Chen, S.-L. (2021). Electronegative very-low-density lipoprotein induces brain inflammation and cognitive dysfunction in mice. Scientific Reports, 11(1), 6013.

Lofgren, M., & Hinzen, W. (2022). Breaking the flow of thought: increase of empty pauses in the connected speech of people with mild and moderate Alzheimer’s disease. Journal of Communication Disorders, 97, 106214.

Lundberg, S. (2017). A unified approach to interpreting model predictions. arXiv preprint arXiv:1705.07874.

Maheux, E., Koval, I., Ortholand, J., Birkenbihl, C., Archetti, D., Bouteloup, V., Epelbaum, S., Dufouil, C., Hofmann-Apitius, M., & Durrleman, S. (2023). Forecasting individual progression trajectories in Alzheimer’s disease. Nature Communications, 14(1), 761.

Meulenbroek, O., Rijpkema, M., Kessels, R. P., Rikkert, M. G. O., & Fernández, G. (2010). Autobiographical memory retrieval in patients with Alzheimer’s disease. NeuroImage, 53(1), 331–340.

Mirabnahrazam, G., Ma, D., Beaulac, C., Lee, S., Popuri, K., Lee, H., Cao, J., Galvin, J. E., Wang, L., & Beg, M. F. (2023). Predicting time-to-conversion for dementia of Alzheimer’s type using multi-modal deep survival analysis. Neurobiology of Aging, 121, 139–156.

Mo, T., Lam, J. C. K., Li, V. O. K., & Cheung, L. Y. L. (2024). Leveraging Large Language Models for Identifying Interpretable Linguistic Markers and Enhancing Alzheimer’s Disease Diagnostics. medRxiv, 2024.2008. 2022.24312463.

Moor, M., Banerjee, O., Abad, Z. S. H., Krumholz, H. M., Leskovec, J., Topol, E. J., & Rajpurkar, P. (2023). Foundation models for generalist medical artificial intelligence. Nature, 616(7956), 259–265.

Mueller, K. D., Hermann, B., Mecollari, J., & Turkstra, L. S. (2018). Connected speech and language in mild cognitive impairment and Alzheimer’s disease: A review of picture description tasks. Journal of Clinical and Experimental Neuropsychology, 40(9), 917–939.

Nguyen, H. H., Blaschko, M. B., Saarakkala, S., & Tiulpin, A. (2023). Clinically-inspired multi-agent transformers for disease trajectory forecasting from multimodal data. IEEE Transactions on Medical Imaging.

OpenAI. (n.d.-a). Introducing Whisper. Retrieved 15 Sep 2024 from https://openai.com/index/whisper/

OpenAI. (n.d.-b). OpenAI’s GPT-4o. Retrieved 15 Sep 2024 from https://platform.openai.com/docs/models/gpt-4o

Pereira, J. B., Mijalkov, M., Kakaei, E., Mecocci, P., Vellas, B., Tsolaki, M., Kłoszewska, I., Soininen, H., Spenger, C., & Lovestone, S. (2016). Disrupted network topology in patients with stable and progressive mild cognitive impairment and Alzheimer’s disease. Cerebral Cortex, 26(8), 3476–3493.

Pezzoli, S., Giorgio, J., Martersteck, A., Dobyns, L., Harrison, T. M., & Jagust, W. J. (2024). Successful cognitive aging is associated with thicker anterior cingulate cortex and lower tau deposition compared to typical aging. Alzheimer’s & Dementia, 20(1), 341–355.

Qian, X.-h., Xie, R.-y., Liu, X.-l., & Tang, H.-d. (2022). Mechanisms of short-chain fatty acids derived from gut microbiota in Alzheimer’s disease. Aging and disease, 13(4), 1252.

Rousseeuw, P. J. (1987). Silhouettes: a graphical aid to the interpretation and validation of cluster analysis. Journal of Computational and Applied Mathematics, 20, 53–65.

Sama, D. M., & Norris, C. M. (2013). Calcium dysregulation and neuroinflammation: discrete and integrated mechanisms for age-related synaptic dysfunction. Ageing Research Reviews, 12(4), 982–995.

Seedat, N., Huynh, N., van Breugel, B., & van der Schaar, M. (2023). Curated LLM: Synergy of LLMs and data curation for tabular augmentation in ultra low-data regimes. arXiv preprint arXiv:2312.12112.

Simonyan, K. (2013). Deep inside convolutional networks: Visualising image classification models and saliency maps. arXiv preprint arXiv:1312.6034.

Singhal, K., Azizi, S., Tu, T., Mahdavi, S. S., Wei, J., Chung, H. W., Scales, N., Tanwani, A., Cole-Lewis, H., & Pfohl, S. (2023). Large language models encode clinical knowledge. Nature, 620(7972), 172–180.

Stefano, P., Concetta, C., Luigi, D., Marco, C., Antonis, P., Ioannis, L., Diego, A., Rita, A. A., Raffaele, S., & Ilaria, R. (2016). Role of neurodevelopment involved genes in psychiatric comorbidities and modulation of inflammatory processes in Alzheimer’s disease. Journal of the Neurological Sciences, 370, 162–166.

Thirunavukarasu, A. J., Ting, D. S. J., Elangovan, K., Gutierrez, L., Tan, T. F., & Ting, D. S. W. (2023). Large language models in medicine. Nature Medicine, 29(8), 1930–1940.

Wang, C., Li, Y., Tsuboshita, Y., Sakurai, T., Goto, T., Yamaguchi, H., Yamashita, Y., Sekiguchi, A., Tachimori, H., & Alzheimer’s Disease Neuroimaging Initiative. (2022). A high-generalizability machine learning framework for predicting the progression of Alzheimer’s disease using limited data. npj Digital Medicine, 5(1), 43.

Whiley, L., Sen, A., Heaton, J., Proitsi, P., García-Gómez, D., Leung, R., Smith, N., Thambisetty, M., Kloszewska, I., & Mecocci, P. (2014). Evidence of altered phosphatidylcholine metabolism in Alzheimer’s disease. Neurobiology of Aging, 35(2), 271–278.

Xu, L., Wu, H., He, C., Wang, J., Zhang, C., Nie, F., & Chen, L. (2022). Multi-modal sequence learning for Alzheimer’s disease progression prediction with incomplete variable-length longitudinal data. Medical Image Analysis, 82, 102643.

Yi, F., Zhang, Y., Yuan, J., Liu, Z., Zhai, F., Hao, A., Wu, F., Somekh, J., Peleg, M., & Zhu, Y.-C. (2023). Identifying underlying patterns in Alzheimer’s disease trajectory: a deep learning approach and Mendelian randomization analysis. EClinicalMedicine, 64.

Young, A. L., Oxtoby, N. P., Garbarino, S., Fox, N. C., Barkhof, F., Schott, J. M., & Alexander, D. C. (2024). Data-driven modelling of neurodegenerative disease progression: thinking outside the black box. Nature Reviews Neuroscience, 25(2), 111–130.

Yuan, C., Linn, K. A., & Hubbard, R. A. (2023). Algorithmic Fairness of Machine Learning Models for Alzheimer Disease Progression. JAMA Network Open, 6(11), e2342203–e2342203.

Zhang, J., Chen, L., Ye, Y., Guo, G., Chen, R., Vanasse, A., & Wang, S. (2020). Survival neural networks for time-to-event prediction in longitudinal study. Knowledge and Information Systems, 62, 3727–3751.

Zhang, Q., Han, Y., Lam, J. C. K., Bai, R., Gozes, I., & Li, V. O. K. (2023). An Expert-guided Hierarchical Graph Attention Network for Post-traumatic Stress Disorder Highly-associative Genetic Biomarkers Identification. medRxiv, 2023.2001. 2030.23285175.

